# Tocilizumab in patients admitted to hospital with COVID-19 (RECOVERY): preliminary results of a randomised, controlled, open-label, platform trial

**DOI:** 10.1101/2021.02.11.21249258

**Authors:** RECOVERY Collaborative Group, Peter W Horby, Guilherme Pessoa-Amorim, Leon Peto, Christopher E Brightling, Rahuldeb Sarkar, Koshy Thomas, Vandana Jeebun, Abdul Ashish, Redmond Tully, David Chadwick, Muhammad Sharafat, Richard Stewart, Banu Rudran, J Kenneth Baillie, Maya H Buch, Lucy C Chappell, Jeremy N Day, Saul N Furst, Thomas Jaki, Katie Jeffery, Edmund Juszczak, Wei Shen Lim, Alan Montgomery, Andrew Mumford, Kathryn Rowan, Guy Thwaites, Marion Mafham, Richard Haynes, Martin J Landray

## Abstract

**Background:** Tocilizumab is a monoclonal antibody that binds to the receptor for interleukin (IL)-6, reducing inflammation, and is commonly used to treat rheumatoid arthritis. We evaluated the safety and efficacy of tocilizumab in adult patients admitted to hospital with COVID-19 with evidence of both hypoxia and systemic inflammation.

**Methods:** This randomised, controlled, open-label, platform trial (Randomised Evaluation of COVID-19 Therapy [RECOVERY]), is assessing several possible treatments in patients hospitalised with COVID-19 in the UK. Those trial participants with hypoxia (oxygen saturation <92% on air or requiring oxygen therapy) and evidence of systemic inflammation (C-reactive protein [CRP] ≥75 mg/L) were eligible for randomisation to usual standard of care alone versus usual standard of care plus tocilizumab at a dose of 400 mg to 800 mg (depending on weight) given intravenously. A second dose could be given 12 to 24 hours later if the patient’s condition had not improved. The primary outcome was 28-day mortality, assessed in the intention-to-treat population. The trial is registered with ISRCTN (50189673) and clinicaltrials.gov (NCT04381936).

**Findings:** Between 23 April 2020 and 24 January 2021, 4116 adults were included in the assessment of tocilizumab, including 562 (14%) patients receiving invasive mechanical ventilation, 1686 (41%) receiving non-invasive respiratory support, and 1868 (45%) receiving no respiratory support other than oxygen. Median CRP was 143 [IQR 107-204] mg/L and 3385 (82%) patients were receiving systemic corticosteroids at randomisation. Overall, 596 (29%) of the 2022 patients allocated tocilizumab and 694 (33%) of the 2094 patients allocated to usual care died within 28 days (rate ratio 0·86; 95% confidence interval [CI] 0·77-0·96; p=0·007). Consistent results were seen in all pre-specified subgroups of patients. In particular, a clear mortality benefit was seen in those receiving systemic corticosteroids. Patients allocated to tocilizumab were more likely to be discharged from hospital alive within 28 days (54% vs. 47%; rate ratio 1·22; 95% CI 1·12-1·34; p<0·0001). Among those not receiving invasive mechanical ventilation at baseline, patients allocated tocilizumab were less likely to reach the composite endpoint of invasive mechanical ventilation or death (33% vs. 38%; risk ratio 0·85; 95% CI 0·78-0·93; p=0·0005).

**Interpretation:** In hospitalised COVID-19 patients with hypoxia and systemic inflammation, tocilizumab improved survival and other clinical outcomes. These benefits were seen regardless of the level of respiratory support and were additional to the benefits of systemic corticosteroids.

**Funding:** UK Research and Innovation (Medical Research Council) and National Institute of Health Research (Grant ref: MC_PC_19056).

## INTRODUCTION

The majority of severe acute respiratory syndrome coronavirus 2 (SARS-CoV-2) infections are either asymptomatic or result in only mild disease.^1^ However, a substantial proportion of infected individuals develop a respiratory illness requiring hospital care, which can progress to critical illness with hypoxic respiratory failure requiring prolonged ventilatory support. Among COVID-19 patients admitted to UK hospitals in Spring 2020, the case fatality rate was over 26%, and was in excess of 37% in patients requiring invasive mechanical ventilation.^2^

Hypoxic respiratory failure in patients with COVID-19 is associated with evidence of systemic inflammation, including release of pro-inflammatory cytokines, such as interleukin (IL)-1, IL-6 and TNFα, and elevated levels of D-dimers, ferritin, and C-reactive protein (CRP).^3,4^ The host immune response is thought to play a key role in driving an acute inflammatory pneumonic process with diffuse alveolar damage, myeloid cell infiltrates, and microvascular thrombosis.^5^ The beneficial effects of dexamethasone and other corticosteroids in COVID-19 patients with hypoxic lung damage suggest that other, more specific immunomodulatory agents may provide additional improvements in clinical outcomes.^6,7^

Tocilizumab is a recombinant humanised anti-IL-6 receptor monoclonal antibody that inhibits the binding of IL-6 to both membrane and soluble IL-6 receptors, blocking IL-6 signalling and reducing inflammation. Tocilizumab is licensed in the UK as an intravenous treatment for patients with rheumatoid arthritis and for people with chimeric antigen receptor T cell-induced severe or life-threatening cytokine release syndrome.

Randomised trials of tocilizumab in COVID-19 have so far shown mixed results for 28-day mortality: six small trials reported no benefit while the somewhat larger REMAP-CAP trial reported a benefit in patients requiring organ support.^8-14^ Here we report the results of a large randomised controlled trial of tocilizumab in adult patients hospitalised with severe COVID-19 characterised by hypoxia and significant inflammation.

## METHODS

### Study design and participants

The Randomised Evaluation of COVID-19 therapy (RECOVERY) trial is an investigator-initiated, individually randomised, controlled, open-label, platform trial to evaluate the effects of potential treatments in patients hospitalised with COVID-19. Details of the trial design and results for other possible treatments have been published previously.^6,15-17^ The trial is being conducted in acute National Health Service (NHS) hospitals in the UK, supported by the National Institute for Health Research Clinical Research Network. The trial is coordinated by the Nuffield Department of Population Health at University of Oxford (Oxford, UK), the trial sponsor. The trial is conducted in accordance with the principles of the International Conference on Harmonisation–Good Clinical Practice guidelines and approved by the UK Medicines and Healthcare products Regulatory Agency (MHRA) and the Cambridge East Research Ethics Committee (ref: 20/EE/0101). The protocol, statistical analysis plan, and additional information are available on the study website www.recoverytrial.net. This report is limited to adult patients. The randomised assessment of tocilizumab in children under 18 years old is ongoing.

Patients admitted to hospital were eligible for the study if they had clinically suspected or laboratory confirmed SARS-CoV-2 infection and no medical history that might, in the opinion of the attending clinician, put the patient at significant risk if they were to participate in the trial. Written informed consent was obtained from all patients, or their legal representative if they were too unwell or unable to provide consent.

### Randomisation and masking

Data were collected at study entry using a web-based case report form that included demographics and major comorbidities (appendix p 32). All eligible and consenting patients received usual standard of care and underwent an initial (main) randomisation comprising up to 3 parts in a factorial design (appendix p 29): part 1, no additional treatment vs. either dexamethasone, lopinavir-ritonavir, hydroxychloroquine, azithromycin, or colchicine; part 2, no additional treatment vs. either convalescent plasma or REGN-COV2 (a combination of two monoclonal antibodies directed against SARS-CoV-2 spike protein); and part 3, no additional treatment vs. aspirin. Over time, treatment arms were added to and removed from the protocol (appendix p 26), and not all treatments were available at every hospital. Similarly, not all treatments were suitable for some patients (e.g. due to comorbid conditions or concomitant medication). In any of these cases, randomisation was between fewer arms.

Up to 21 days after the main randomisation and regardless of treatment allocation, RECOVERY trial participants with clinical evidence of progressive COVID-19 (defined as oxygen saturation <92% on room air or receiving oxygen therapy, and CRP ≥75 mg/L) could be considered for randomisation to tocilizumab vs. usual care alone. Baseline data collected for this second randomisation included level of respiratory support, markers of progressive COVID-19 (including oxygen saturation, CRP, ferritin, creatinine), suitability for the study treatment, and treatment availability at the site (appendix p 34). For some patients, tocilizumab was unavailable at the hospital at the time of enrolment or was considered by the managing physician to be either definitely indicated or definitely contraindicated. In such cases, the patients were not eligible for the tocilizumab randomisation. Patients with known hypersensitivity to tocilizumab, evidence of active tuberculosis infection or clear evidence of active bacterial, fungal, viral, or other infection (besides COVID-19) were not eligible for randomisation to tocilizumab.

Patients who were eligible for randomisation to tocilizumab were assigned to either usual standard of care or usual standard of care plus tocilizumab in a 1:1 ratio using web-based simple (unstratified) randomisation with allocation concealed until after randomisation. Patients allocated to tocilizumab were to receive tocilizumab as a single intravenous infusion over 60 minutes. The dose of tocilizumab was determined by body weight (800 mg if weight >90kg; 600 mg if weight >65 and ≤90 kg; 400 mg if weight >40 and ≤65 kg; and 8mg/kg if weight ≤40 kg). A second dose could be given 12 to 24 hours later if, in the opinion of the attending clinician, the patient’s condition had not improved. Allocated treatment was prescribed by the managing doctor. Roche Products Ltd (UK) supported the trial through provision of tocilizumab. Participants and local study staff were not masked to the allocated treatment. The steering committee, investigators, and all others involved in the trial were masked to the outcome data during the trial.

### Procedures

A single online follow-up form was completed when participants were discharged, had died or at 28 days after the initial randomisation, whichever occurred earliest (appendix p 36-41). Information was recorded on adherence to allocated study treatment, receipt of other COVID-19 treatments, duration of admission, receipt of respiratory or renal support, and vital status (including cause of death). In addition, routine healthcare and registry data were obtained for the full follow-up period, including information on vital status (with date and cause of death), discharge from hospital, receipt of respiratory support, or renal replacement therapy.

### Outcomes

Outcomes were assessed at 28 days after randomisation to tocilizumab vs. usual care alone, with further analyses specified at 6 months. The primary outcome was all-cause mortality. Secondary outcomes were time to discharge alive from hospital, and, among patients not receiving invasive mechanical ventilation at randomisation, receipt of invasive mechanical ventilation (including extra-corporeal membrane oxygenation) or death. Prespecified subsidiary clinical outcomes were use of non-invasive respiratory support (defined as high flow nasal oxygen, continuous positive airway pressure, or non-invasive ventilation), time to successful cessation of invasive mechanical ventilation (defined as cessation of invasive mechanical ventilation within, and survival to, 28 days), and use of renal dialysis or haemofiltration. Prespecified safety outcomes included cause-specific mortality and major cardiac arrhythmia. Information on suspected serious adverse reactions was collected in an expedited fashion to comply with regulatory requirements.

### Statistical Analysis

In accordance with the statistical analysis plan (version 2.1 appendix pp 93-117), an intention-to-treat comparison was conducted between patients who entered the randomised comparison of tocilizumab vs. usual care. For the primary outcome of 28-day mortality, the log-rank observed minus expected statistic and its variance were used to test the null hypothesis of equal survival curves (i.e., the log-rank test) and to calculate the one-step estimate of the average mortality rate ratio. We constructed Kaplan-Meier survival curves to display cumulative mortality over the 28-day period. For this preliminary report, information on the primary outcome is available for 92% of randomised patients. Those patients who had not been followed for 28 days and were not known to have died by the time of the data cut for this <u>preliminary</u> analysis (8 February 2021) were either censored on 8 February 2021 or, if they had already been discharged alive, were right-censored for mortality at day 29 (that is, in the absence of any information to the contrary they were assumed to have survived 28 days). [Note: This censoring rule will not be necessary when all patients have completed the 28 day follow-up period on 25 February 2021.] We used the same method to analyse time to hospital discharge and successful cessation of invasive mechanical ventilation, with patients who died in hospital right-censored on day 29. For the pre-specified composite secondary outcome of invasive mechanical ventilation or death within 28 days (among those not receiving invasive mechanical ventilation at randomisation) and the subsidiary clinical outcomes of receipt of ventilation and receipt of haemodialysis or haemofiltration, the precise dates were not available and so the risk ratio was estimated instead.

Prespecified analyses of the primary outcome were performed in subgroups defined by six characteristics at the time of randomisation: age, sex, ethnicity, level of respiratory support, days since symptom onset, and use of systemic corticosteroids (including dexamethasone). Observed effects within subgroup categories were compared using a chi-squared test for heterogeneity or trend, in accordance with the prespecified analysis plan.

Estimates of rate and risk ratios are shown with 95% confidence intervals. All p-values are 2-sided and are shown without adjustment for multiple testing. The full database is held by the study team which collected the data from study sites and performed the analyses at the Nuffield Department of Population Health, University of Oxford (Oxford, UK).

Prior to commencement of the randomisation to tocilizumab vs. usual care, the trial steering committee determined that if 28-day mortality in the usual care group was above 25% then recruitment of around 4000 patients to this comparison would provide 90% power at two-sided P=0.01 to detect a proportional reduction in 28-day mortality of one-fifth. Consequently, Roche Products Ltd provided sufficient treatment for 2000 patients to receive tocilizumab. The trial steering committee, masked to the results, closed recruitment to the tocilizumab comparison at the end of January 24, 2021 as over 4000 patients had been randomised.

Analyses were performed using SAS version 9.4 and R version 3.4. The trial is registered with ISRCTN (50189673) and clinicaltrials.gov (NCT04381936).

## RESULTS

Between 14 April 2020 and 24 January 2021, 4116 (19%) of 21550 patients enrolled into the RECOVERY trial at one of the 131 sites in the UK participating in the tocilizumab comparison were eligible for randomisation. 2022 patients were randomly allocated to tocilizumab and 2094 were randomly allocated to usual care. The mean age of these participants was 63·6 years (SD 13·7). At randomisation, 562 (14%) patients were receiving invasive mechanical ventilation, 1686 (41%) were receiving non-invasive respiratory support (including high-flow nasal oxygen, continuous positive airway pressure, and non-invasive ventilation), and 1868 (45%) were receiving no respiratory support other than simple oxygen therapy (9 of these patients were reportedly not receiving oxygen at randomisation) (table 1). Median CRP was 143 [IQR 107-204] mg/L. 82% of patients were reported to be receiving corticosteroids at randomisation (and 97% of the patients enrolled since the announcement of the dexamethasone result from RECOVERY in June 2021).

**Table 1:** Baseline characteristics by randomised allocation.

|  | **Tocilizumab**<br>**(n=2022)** | **Usual care**<br>**(n=2094)** |
| --- | --- | --- |
| Mean (SD) Age, years | 63.3 (13.7) | 63.9 (13.6) |
| ≥18 to <70 | 1332 (66%) | 1354 (65%) |
| ≥70 to <80 | 477 (24%) | 480 (23%) |
| ≥80 | 213 (11%) | 260 (12%) |
| Sex |  |  |
| Male | 1335 (66%) | 1437 (69%) |
| Female* | 687 (34%) | 657 (31%) |
| Ethnicity |  |  |
| White | 1356 (67%) | 1426 (68%) |
| Black, Asian, or Minority Ethnic | 341 (17%) | 357 (17%) |
| Unknown | 325 (16%) | 311 (15%) |
| Number of days since symptom onset | 9 (7-13) | 10 (7-14) |
| Number of days since hospitalisation | 2 (1-5) | 2 (1-5) |
| Oxygen saturation, % | 94 (92-96) | 94 (91-95) |
| Respiratory support at second randomisation |  |  |
| No ventilator support† | 935 (46%) | 933 (45%) |
| Non-invasive ventilation‡ | 819 (41%) | 867 (41%) |
| Invasive mechanical ventilation§ | 268 (13%) | 294 (14%) |
| Biochemistry at second randomisation |  |  |
| Latest C-reactive protein, mg/L | 143 (107-203) | 144 (106-205) |
| Ferritin, ng/mL | 947 (497-1599) | 944 (507-1533) |
| Creatinine, umol/L | 77 (62-98) | 77 (62-100) |
| Previous diseases |  |  |
| Diabetes | 569 (28%) | 600 (29%) |
| Heart disease | 435 (22%) | 497 (24%) |
| Chronic lung disease | 473 (23%) | 484 (23%) |
| Tuberculosis | 3 (<1%) | 5 (<1%) |
| HIV | 7 (<1%) | 8 (<1%) |
| Severe liver disease¶ | 14 (<1%) | 10 (<1%) |
| Severe kidney impairment\|\| | 118 (6%) | 99 (5%) |
| Any of the above | 1100 (54%) | 1163 (56%) |
| SARS-Cov-2 test result |  |  |
| Positive | 1891 (94%) | 1967 (94%) |
| Negative | 68 (3%) | 66 (3%) |
| Test result not yet known | 63 (3%) | 61 (3%) |
| First randomisation\$ | | |
| Number of days since first randomisation | 0 (0-1) | 0 (0-1) |

Tocilizumab for COVID-19
|  | **Tocilizumab**<br>**(n=2022)** | **Usual care**<br>**(n=2094)** |
| --- | --- | --- |
| Part A allocation |  |  |
| Usual care | 839 (41%) | 869 (41%) |
| Lopinavir/ritonavir | 51 (3%) | 64 (3%) |
| Dexamethasone | 49 (2%) | 45 (2%) |
| Hydroxychloroquine | 37 (2%) | 38 (2%) |
| Azithromycin | 197 (10%) | 177 (8%) |
| Use of systemic corticosteroids<sup>^</sup> |  |  |
| Yes | 1664 (82%) | 1721 (82%) |
| No | 357 (18%) | 367 (18%) |
| Unknown | 1 (<1%) | 6 (<1%) |

Data are mean (SD), n(%), or median (IQR). Information on sex, ethnicity, and SARS-CoV-2 test result were recorded on the main randomisation form when patients first entered the study. All other information was recorded on the second randomisation form (when patients were randomly assigned to tocilizumab vs. usual care alone). \* includes 3 pregnant women. † Includes 9 patients not receiving any oxygen and 1859 patients receiving low-flow oxygen. ‡ includes patients receiving high-flow nasal oxygen, continuous positive airway pressure, or other non-invasive ventilation). § Includes patients receiving invasive mechanical ventilation or extra-corporeal membranous oxygenation. ¶ Defined as requiring ongoing specialist care. || Defined as estimated glomerular filtration rate <30 mL/min/1.73m<sup>2</sup> § 2631 and 1615 participants were randomised into parts B and C of the first randomisation respectively. ^Information on use of corticosteroids was collected from 18 June 2020 onwards following announcement of the results of the dexamethasone comparison from the RECOVERY trial. Participants undergoing first randomisation prior to this date (and who were not allocated to dexamethasone) are assumed not to be receiving systemic corticosteroids.

The follow-up form was completed for 1602 (79%) of 2022 randomised patients in the tocilizumab group and 1664 (79%) of 2094 patients in the usual care group. [Follow-up forms are expected for >95% of participants by the time of the final analyses.] Among patients with a completed follow-up form, 1333 (83%) allocated to tocilizumab and 44 (3%) of those allocated to usual care received at least one dose of tocilizumab (or sarilumab, another IL-6 antagonist; figure 1; webtable 1). 461 (29%) patients in the tocilizumab group and 10 (<1%) in the usual care group received more than 1 dose of tocilizumab (or sarilumab). Use of other treatments for COVID-19 during the 28 days after randomisation was similar among patients allocated tocilizumab and among those allocated usual care (webtable 1).

**Figure 1:**
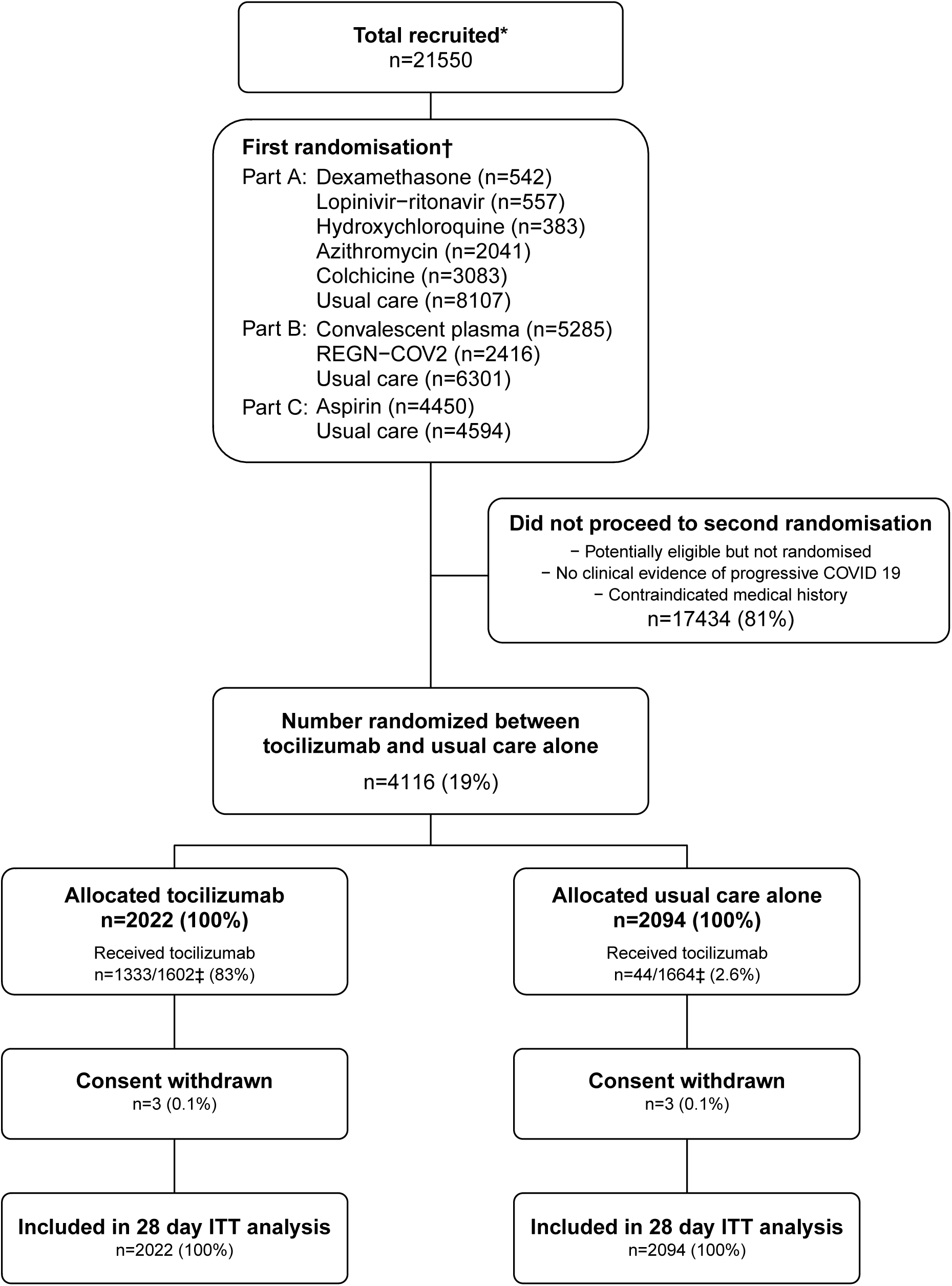
Trial profile – Flow of participants through the RECOVERY trial. ITT=intention to treat. *Number of adult patients recruited at a site activated for the tocilizumab comparison. †The first randomisation comprised up to 3 factorial elements such that an eligible patient could be entered into between 1 and 3 randomised comparisons, depending on the then current protocol, the patients suitability for particular treatments, and the availability of the treatment at the site. ‡ 1602/2022 (79%) patients of those allocated to tocilizumab and 1664/2094 (79%) of those allocated to usual care had a completed follow–up form at time of analysis.

Allocation to tocilizumab was associated with a significant reduction in the primary outcome of 28-day mortality compared with usual care alone (596 [29%] of 2022 patients in the tocilizumab group vs. 694 (33%) of 2094 patients in the usual care group; rate ratio 0·86; 95% confidence interval [CI], 0·77 to 0·96; p=0·007; figure 2a). In an exploratory analysis restricted to the 3858 (94%) patients with a positive SARS-CoV-2 test result, the result was similar (rate ratio 0.87, 95% CI 0·78 to 0.98; p=0·02).

**Figure 2:**
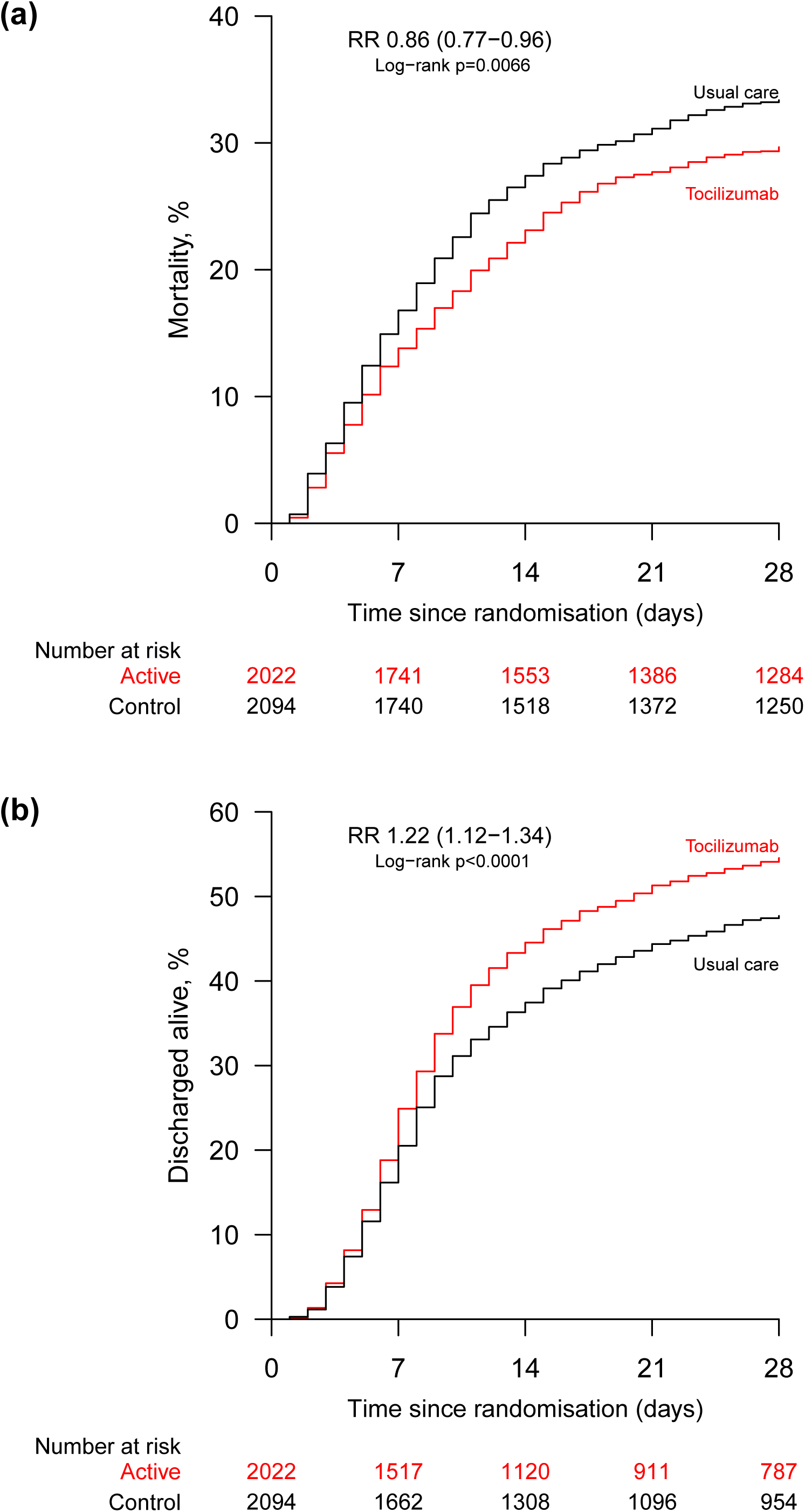
Effect of allocation to tocilizumab on (a) 28–day mortality and (b) discharge from hospital alive within 28 days of randomisation.

Allocation to tocilizumab was associated with a greater probability of discharge from hospital alive within 28 days (54% vs. 47%; rate ratio 1·22, 95% CI 1·12 to 1·34, p<0·0001; figure 2b and table 2). Among those not on invasive mechanical ventilation at baseline, allocation to tocilizumab was associated with a reduction in the risk of progressing to the pre-specified composite secondary outcome of invasive mechanical ventilation or death when compare to usual care alone (33% vs. 38%, risk ratio 0·85, 95% CI 0·78 to 0·93, p=0·0005; table 2).

**Table 2:** Effect of allocation to tocilizumab on main study outcomes.

|  | Treatment allocation |  | RR (95% CI) | p value |
| --- | --- | --- | --- | --- |
|  | Tocilizumab<br>(n=2022) | Usual care<br>(n=2094) |  |  |
| Primary outcome |  |  |  |  |
| Total: 28-day mortality | 596 (29%) | 694 (33%) | 0.86 (0.77-0.96) | 0.0066 |
| Secondary outcomes |  |  |  |  |
| Median time to being discharged alive, days | 20 | >28 |  |  |
| Discharged alive from hospital within 28 days | 1093 (54%) | 990 (47%) | 1.22 (1.12-1.34) | <0.0001 |
| Receipt of invasive mechanical ventilation or death* | 571/1754 (33%) | 687/1800 (38%) | 0.85 (0.78-0.93) | 0.0005 |
| Invasive mechanical ventilation | 215/1754 (12%) | 273/1800 (15%) | 0.81 (0.68-0.95) | 0.01 |
| Death | 471/1754 (27%) | 552/1800 (31%) | 0.88 (0.79-0.97) | 0.01 |
| Subsidiary clinical outcomes |  |  |  |  |
| Receipt of ventilation† | 233/935 (25%) | 242/933 (26%) | 0.96 (0.82-1.12) | 0.61 |
| Non-invasive ventilation | 222/935 (24%) | 223/933 (24%) | 0.99 (0.84-1.17) | 0.94 |
| Invasive mechanical ventilation | 45/935 (5%) | 63/933 (7%) | 0.71 (0.49-1.03) | 0.07 |
| Successful cessation of invasive mechanical ventilation‡ | 91/268 (34%) | 94/294 (32%) | 1.07 (0.80-1.43) | 0.64 |
| Use of haemodialysis or haemofiltration§ | 103/2003 (5%) | 142/2075 (7%) | 0.75 (0.59-0.96) | 0.02 |

Data are n(%), n/N (%), or median (interquartile range). RR=rate ratio for the outcomes of 28-day mortality, hospital discharge and successful cessation of invasive mechanical ventilation, and risk ratio for other outcomes. \* Analyses include only those on no ventilator support or non-invasive ventilation at second randomisation. † Analyses include only those on no ventilator support at second randomisation. ‡ Analyses restricted to those on invasive mechanical ventilation at second randomisation. § Analyses exclude those on haemodialysis or haemofiltration at second randomisation.

We observed similar results across all pre-specified sub-groups (figure 3 and webfigures 2 and 3), including the level of respiratory support at randomisation (figure 3). Given the number of hypothesis tests conducted, the suggestion of a larger proportional mortality reduction among those receiving a corticosteroid compared with those not [interaction p=0.01] may reflect the play of chance. An exploratory analysis showed that the effects of tocilizumab on 28-day mortality were similar for those randomised ≤2 or >2 days since hospitalisation (interaction p=0.86).

**Figure 3:**
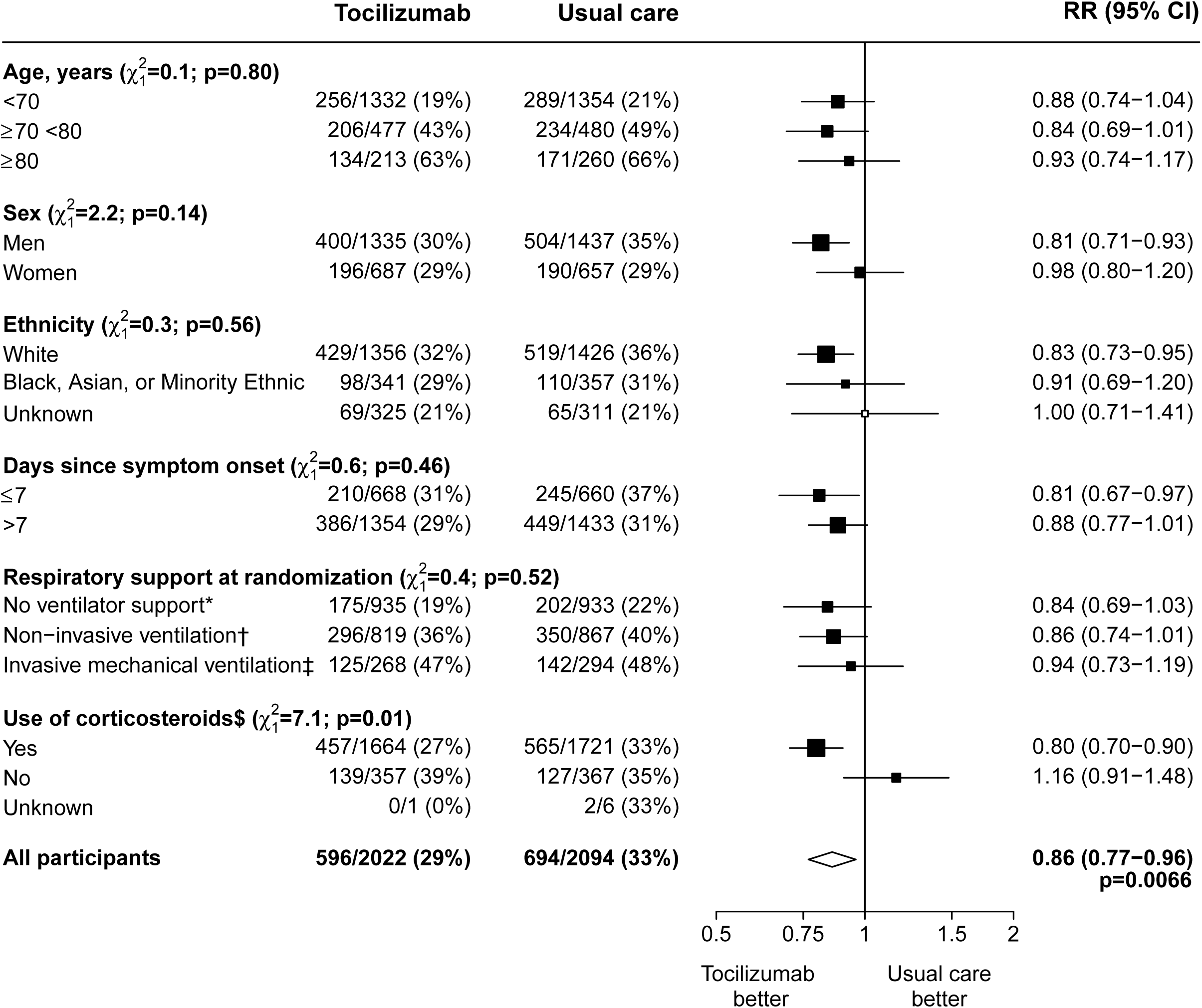
Effect of allocation to tocilizumab on 28–day mortality by baseline characteristics. Subgroup–specific rate ratio estimates are represented by squares (with areas of the squares proportional to the amount of statistical information) and the lines through them correspond to the 95% CIs. The ethnicity and days since onset subgroups exclude those with missing data, but these patients are included in the overall summary diamond. * Includes 9 patients not receiving any oxygen and 1859 patients receiving simple oxygen only. † Includes patients receiving high–flow nasal oxygen, continuous positive airway pressure ventilation, other non–invasive ventilation. ‡ Includes patients receiving invasive mechanical ventilation and extra–corporeal membranous oxygenation. $ Information on use of corticosteroids was collected from 18 June 2020 onwards following announcement of the results of the dexamethasone comparison from the RECOVERY trial. Participants undergoing first randomisation prior to this date (and who were not allocated to dexamethasone) are assumed not to be receiving systemic corticosteroids.

In pre-specified subsidiary analyses, we found no significant effect of tocilizumab on subsequent receipt of non-invasive respiratory support or invasive mechanical ventilation among those on no respiratory support at randomisation (table 2, webfigure 1). Nor was there a significant effect on the rate of successful cessation of invasive mechanical ventilation among those on invasive mechanical ventilation at randomisation. However, allocation to tocilizumab reduced the use of haemodialysis or haemofiltration (5% vs. 7%, risk ratio 0·75, 95% CI 0·59 to 0·96, p=0·02; table 2). Preliminary information on cause-specific mortality shows no evidence of excess deaths from other infections (webtable 2). We observed no significant differences in the frequency of new cardiac arrhythmias (webtable 3). There were three reports of serious adverse reactions believed to be related to tocilizumab: one each of otitis externa, Staphylococcus aureus bacteraemia, and lung abscess, all of which resolved with standard treatment.

## DISCUSSION

The results of this large, randomised trial indicate that tocilizumab is an effective treatment for hospitalised COVID-19 patients who have hypoxia and evidence of inflammation (CRP ≥75 mg/L). Treatment with tocilizumab improved survival and the chances of discharge from hospital alive by 28 days, and reduced the chances of progressing to require invasive mechanical ventilation. These benefits were consistent across all patient groups studied, including those receiving invasive mechanical ventilation, non-invasive respiratory support, or no respiratory support other than simple oxygen. The benefits of tocilizumab were clearly seen among those also receiving treatment with a systemic corticosteroid such as dexamethasone. There was also a significant reduction in the need for haemodialysis or haemofiltration. Although we did not see any effect on the duration of invasive mechanical ventilation, only 185 patients had such ventilation successfully removed within the first 28 days, so power to detect any benefit was low.

Since mid-2020, seven randomised controlled trials of tocilizumab for the treatment of COVID-19 have reported. These include six small trials (fewer than 100 deaths in each) and the somewhat larger REMAP-CAP trial, which recruited critically ill patients with COVID-19, over 99% of whom required non-invasive respiratory support or invasive mechanical ventilation.^8-14^ Taken together, these previous trials did not show a significant mortality benefit for treatment with tocilizumab (death rate ratio 0·91, 95% CI 0·72-1·14, figure 4). The RECOVERY trial contains over three times as many deaths as all the previous trials combined. When all 8 trials are considered together, allocation to tocilizumab is associated with a 13% proportional reduction in 28-day mortality (death rate ratio 0·87, 95% CI 0·79-0·96, p=0·005, figure 4).

**Figure 4:**
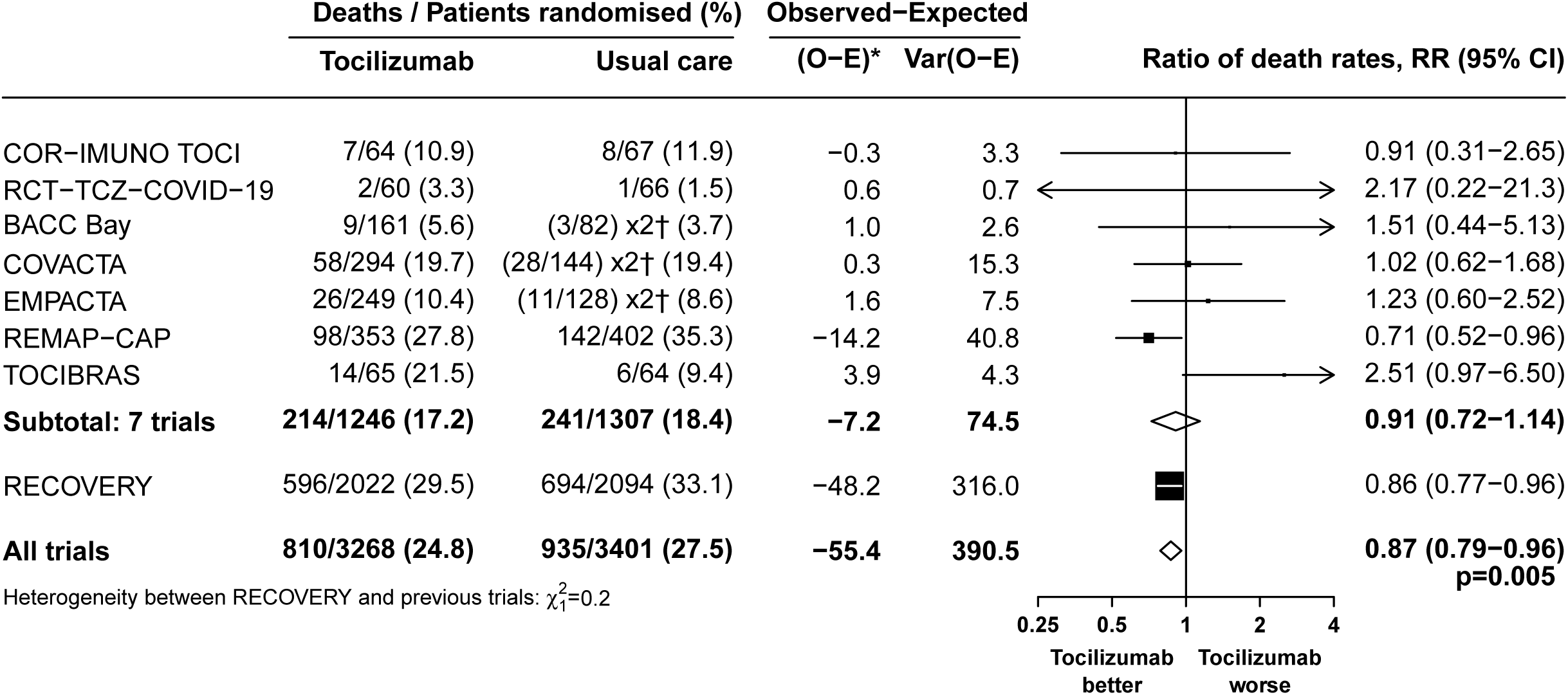
Tocilizumab vs usual care in patients hospitalised with COVID – Meta–analysis of mortality in RECOVERY and other trials. * Log–rank O–E for RECOVERY, O–E from 2×2 tables for the other trials. RR is calculated by taking ln RR to be (O–E)/V with Normal variance 1/V. Subtotals or totals of (O–E) and of V yield inverse–variance–weighted averages of the ln RR values. † For balance, controls in the 2:1 studies count twice in the control totals and subtotals.

These benefits are in addition to those previously reported for corticosteroids, which is now usual standard of care for COVID-19 patients requiring treatment with oxygen.^6,18^ Our data suggest that in COVID-19 patients who are hypoxic and have evidence of systematic inflammation, treatment with a combination of a systemic corticosteroid plus tocilizumab would be expected to reduce mortality by about one-third for patients receiving simple oxygen and nearly one-half for those receiving invasive mechanical ventilation.

Previous trials have provided weak evidence that tocilizumab may shorten time to discharge or reduce progression to invasive mechanical ventilation or death.^10,14^ Our results show that in a broad spectrum of patients, tocilizumab increases the chances of being discharged alive within 28 days and reduces the chance of progression to receiving invasive mechanical ventilation. As with the mortality benefit, these effects are consistent regardless of the level of respiratory support at the time of enrolment.

Strengths of this trial included that it was randomised, had a large sample size, and included patients requiring various levels of respiratory support (from simple oxygen through to invasive mechanical ventilation). There are some limitations: For this preliminary report, information on the primary outcome is available for 92% of patients. This should increase to >99% by early March when all patients have passed the 28-day follow-up period. Following random assignment, 17% of patients in the tocilizumab group did not receive this treatment. The reasons for this were not recorded. The size of the effects of tocilizumab reported in this paper are therefore an underestimate of the true effects of actually using the treatment. Hospital stay is very long for many of these patients (median >28 days); the pre-planned analyses at 6 months will provide additional information on the full effects of tocilizumab on clinical outcomes.

The RECOVERY results support the use of tocilizumab, but other IL-6 antagonists are available. Although the effects of sarilumab in REMAP-CAP were similar to tocilizumab, only 48 participants received sarilumab.^8^ Two larger trials of sarilumab have completed but have not reported any results (NCT044327388, NTC044315298). Publication of results from those trials is now essential to assess whether alternative IL-6 antagonists to tocilizumab are effective.

Guidelines on the use of IL-6 antagonists for patients with severe COVID-19 vary. For example the US National Institutes for Health conclusion is that there are insufficient data to recommend either for or against the use of tocilizumab or sarilumab, a view shared by some commentators.^19,20^ By contrast, interim guidance in the NHS states that IL-6 antagonists should be considered for patients within 24 hours of starting non-invasive respiratory support or invasive mechanical ventilation.^21^ Our results show that the benefits of tocilizumab extend to a broader group of patients receiving oxygen with or without other forms of respiratory support, and that those benefits include a reduction in the need for invasive mechanical ventilation and other organ support such as renal replacement therapy. Since complicating bacterial infections are infrequent in the early hospitalisation period of COVID-19, this recognised concern in relation to the use of tocilizumab would be lessened with earlier use.^22^

Based upon the ISARIC4C database, approximately 49% of hospitalised COVID-19 patients in the UK would meet our inclusion criteria and hence would benefit from tocilizumab (personal communication, ISARIC4C Investigators). CRP was chosen as the biomarker for inflammation in this study since it is widely used and affordable worldwide, it is correlated with serum IL-6 levels, and early clinical studies of COVID-19 had reported it to be associated with severity and prognosis, with a value of >50 mg/L associated with severe disease and a level of around 75 mg/L distinguishing fatal from non-fatal cases.^23-28^ Whether hypoxic patients with a CRP <75 mg/L could benefit from tocilizumab is unknown. Further work is also needed to consider the health economic benefits of tocilizumab and related IL-6 inhibitors in terms of both patient outcomes and usage of healthcare resources (duration of hospital stay, and frequency of invasive mechanical ventilation and renal replacement therapy).

The RECOVERY trial has demonstrated that for patients hospitalized with severe COVID, treatment with tocilizumab reduces mortality, increases the chances of successful hospital discharge, and reduces the chances of requiring invasive mechanical ventilation. These benefits are additional to those previously reported for dexamethasone. These findings require an update to clinical guidelines and efforts to increase the global availability and affordability of tocilizumab.

## Data Availability

The protocol, consent form, statistical analysis plan, definition & derivation of clinical characteristics & outcomes, training materials, regulatory documents, and other relevant study materials are available online at www.recoverytrial.net. As described in the protocol, the trial Steering Committee will facilitate the use of the study data and approval will not be unreasonably withheld. Deidentified participant data will be made available to bona fide researchers registered with an appropriate institution within 3 months of publication. However, the Steering Committee will need to be satisfied that any proposed publication is of high quality, honours the commitments made to the study participants in the consent documentation and ethical approvals, and is compliant with relevant legal and regulatory requirements (e.g. relating to data protection and privacy). The Steering Committee will have the right to review and comment on any draft manuscripts prior to publication.

https://www.ndph.ox.ac.uk/data-access

## Contributors

This manuscript was initially drafted by the PWH and MJL, further developed by the Writing Committee, and approved by all members of the trial steering committee. PWH and MJL vouch for the data and analyses, and for the fidelity of this report to the study protocol and data analysis plan. PWH, MM, JKB, LCC, SNF, TJ, KJ, WSL, AM, KR, EJ, RH, and MJL designed the trial and study protocol. MM, AR, G P-A, CB, RS, ICT, VJ, AA, RPT, DC, MS, RS, BR, RH, the Data Linkage team at the RECOVERY Coordinating Centre, Health Records, and Local Clinical Centre staff listed in the appendix collected the data. ES, NS, and JRE verified the data and did the statistical analysis. All authors contributed to data interpretation and critical review and revision of the manuscript. PWH and MJL had access to the study data and had final responsibility for the decision to submit for publication.

## Writing Committee (on behalf of the RECOVERY Collaborative Group)

Professor Peter W Horby PhD FRCP,^a,*^ Mark Campbell FRCPath,^b,c,*^ Natalie Staplin PhD,^b,d,*^ Enti Spata,^b,d,*^ Professor Jonathan R Emberson PhD,^b,d^ Guilherme Pessoa-Amorim MD,^b,e^ Leon Peto PhD,^a,c^ Professor Christopher Brightling FRCP,^f^ Rahuldeb Sarkar MPH,^g,h^ Koshy Thomas,^i^ Vandana Jeebun,^j^ Abdul Ashish MD,^k^ Redmond P Tully FRCA,^l^ David Chadwick PhD,^m^ Muhammed Muhammad Sharafat FRCA,^n^ Richard Stewart,° Banu Rudran MRCP,^p^ J Kenneth Baillie MD PhD,^q^ Professor Maya H Buch PhD FRCP,^r^ Professor Lucy C Chappell PhD,^s^ Professor Jeremy N Day PhD FRCP,^a,t^ Professor Saul N Faust PhD FRCPCH,^u^ Professor Thomas Jaki PhD,^v,w^ Katie Jeffery PhD FRCP FRCPath,^c^ Professor Edmund Juszczak MSc,^x^ Professor Wei Shen Lim FRCP,^x,y^ Professor Alan Montgomery PhD,^x^ Professor Andrew Mumford PhD,^z^ Kathryn Rowan PhD,^A^ Professor Guy Thwaites PhD FRCP,^a,t^ Marion Mafham MD,^b,†^ Professor Richard Haynes DM,^b,d,†^ Professor Martin J Landray PhD FRCP.^b,d,e,†^

^a^ Nuffield Department of Medicine, University of Oxford, Oxford, United Kingdom.

^b^ Nuffield Department of Population Health, University of Oxford, Oxford, United Kingdom

^c^ Oxford University Hospitals NHS Foundation Trust, Oxford, United Kingdom

^d^ MRC Population Health Research Unit, University of Oxford, Oxford, United Kingdom

^e^ NIHR Oxford Biomedical Research Centre, Oxford University Hospitals NHS Foundation Trust, Oxford, United Kingdom

^f^ Institute for Lung Health, Leicester NIHR Biomedical Research Centre, University of Leicester, Leicester, United Kingdom

^g^ Medway Foundation NHS Trust, Gillingham, United Kingdom

^h^ King’s College London, London, United Kingdom

^i^ Basildon and Thurrock Hospitals NHS Foundation Trust, Basildon, United Kingdom

^j^ Department of Respiratory Medicine, North Tees & Hartlepool NHS Foundation Trust, Stockton-on-Tees, United Kingdom

^k^ Wrightington Wigan and Leigh NHS Foundation Trust, Wigan, United Kingdom

^l^ Royal Oldham Hospital, Northern Care Alliance, Oldham, United Kingdom

^m^ Centre for Clinical Infection, James Cook University Hospital, Middlesbrough, United Kingdom

^n^ North West Anglia NHS Foundation Trust, Peterborough, United Kingdom

° Milton Keynes University Hospital, Milton Keynes, United Kingdom

^p^ Luton & Dunstable University Hospital, Luton, United Kingdom

^q^ Roslin Institute, University of Edinburgh, Edinburgh, United Kingdom

^r^ Centre for Musculoskeletal Research, University of Manchester, Manchester, and NIHR Manchester Biomedical Research Centre, United Kingdom.

^s^ School of Life Course Sciences, King’s College London, London, United Kingdom

^t^ Oxford University Clinical Research Unit, Ho Chi Minh City, Viet Nam

^u^ NIHR Southampton Clinical Research Facility and Biomedical Research Centre, University Hospital Southampton NHS Foundation Trust and University of Southampton, Southampton, United Kingdom

^v^ Department of Mathematics and Statistics, Lancaster University, Lancaster, United Kingdom

^w^ MRC Biostatistics Unit, University of Cambridge, Cambridge, United Kingdom

^x^ School of Medicine, University of Nottingham, Nottingham, United Kingdom

^y^ Respiratory Medicine Department, Nottingham University Hospitals NHS Trust, Nottingham, United Kingdom

^z^ School of Cellular and Molecular Medicine, University of Bristol, Bristol, United kingdom

^A^ Intensive Care National Audit & Research Centre, London, United Kingdom

*,^†^ equal contribution

## Data Monitoring Committee

Peter Sandercock, Janet Darbyshire, David DeMets, Robert Fowler, David Lalloo, Ian Roberts (until December 2020), M Munavvar (since January 2021), Janet Wittes.

## Data sharing

The protocol, consent form, statistical analysis plan, definition & derivation of clinical characteristics & outcomes, training materials, regulatory documents, and other relevant study materials are available online at www.recoverytrial.net. As described in the protocol, the trial steering committee will facilitate the use of the study data and approval will not be unreasonably withheld. Deidentified participant data will be made available to bona fide researchers registered with an appropriate institution within 3 months of publication. However, the steering committee will need to be satisfied that any proposed publication is of high quality, honours the commitments made to the study participants in the consent documentation and ethical approvals, and is compliant with relevant legal and regulatory requirements (e.g. relating to data protection and privacy). The steering committee will have the right to review and comment on any draft manuscripts prior to publication. Data will be made available in line with the policy and procedures described at: https://www.ndph.ox.ac.uk/data-access. Those wishing to request access should complete the form at https://www.ndph.ox.ac.uk/files/about/data_access_enquiry_form_13_6_2019.docx and e-mailed to

## Acknowledgements

Above all, we would like to thank the thousands of patients who participated in this study. We would also like to thank the many doctors, nurses, pharmacists, other allied health professionals, and research administrators at 176 NHS hospital organisations across the whole of the UK, supported by staff at the National Institute of Health Research (NIHR) Clinical Research Network, NHS DigiTrials, Public Health England, Department of Health & Social Care, the Intensive Care National Audit & Research Centre, Public Health Scotland, National Records Service of Scotland, the Secure Anonymised Information Linkage (SAIL) at University of Swansea, and the NHS in England, Scotland, Wales and Northern Ireland.

The RECOVERY trial is supported by a grant to the University of Oxford from?UK Research and Innovation/NIHR (Grant reference: MC_PC_19056) and by core funding provided by NIHR Oxford Biomedical Research Centre, Wellcome, the Bill and Melinda Gates Foundation, the Department for International Development, Health Data Research UK, the Medical Research Council Population Health Research Unit, the NIHR Health Protection Research Unit in Emerging and Zoonotic Infections, and NIHR Clinical Trials Unit Support Funding. TJ is supported by a grant from UK Medical Research Council (MC_UU_0002/14) and an NIHR Senior Research Fellowship (NIHR-SRF-2015-08-001). WSL is supported by core funding provided by NIHR Nottingham Biomedical Research Centre. Abbvie contributed some supplies of lopinavir-ritonavir for use in this study. Roche Products Ltd supported the trial through the provision of tocilizumab. REGEN-COV2 was provided for this study by Regeneron.

The views expressed in this publication are those of the authors and not necessarily those of the NHS, the National Institute for Health Research, the Department of Health and Social Care, or Roche Products Limited.

